# EHMT2 LOSS-OF-FUNCTION ALTERATIONS CAUSE A KLEEFSTRA-LIKE SYNDROME

**DOI:** 10.1101/2024.01.10.24300997

**Authors:** Beatriz Martinez-Delgado, Estrella Lopez-Martin, Jennifer Kerkhof, Beatriz Baladron, Lidia M. Mielu, Diana Sanchez-Ponce, Ariadna Bada-Navarro, Marina Herrero-Matesanz, Lidia Lopez-Jimenez, Jessica Rzasa, Dmitrijs Rots, Marta Fernandez, Esther Hernandez-San Miguel, Gema Gomez-Mariano, Purificacion Marin-Reina, Rosario Cazorla-Calleja, Javier Alonso, Tjitske Kleefstra, Manuel Posada, Eva Bermejo-Sanchez, Bekim Sadikovic, Maria J. Barrero

## Abstract

Dysregulation of the epigenetic machinery is associated with neurodevelopmental defects in humans. Kleefstra syndrome (KS) is a neurodevelopmental syndrome caused by heterozygous alterations in the gene *EHMT1* that cause loss-of-function. EHTM1 and EHMT2 are highly similar histone methyltransferases that play relevant roles in development. Despite their similarity, individuals with alterations in *EHMT2* have never been described. Here, we describe a pediatric patient with a KS-overlapping phenotype and a single base de novo substitution in *EHMT2* that causes the amino acid change p.Ala1077Ser in the catalytic SET domain. This change causes a reduction in the affinity of the catalytic domain for the H3 tail and in the activity of the enzyme by three- to five-fold. DNA methylation, histone methylation and gene expression profiles suggest a significant overlap between the EHMT2 p.Ala1077Ser variant and KS. Based on this evidence we suggest that EHMT2 haploinsufficiency causes a Kleefstra-like syndrome. Although we cannot rule out dominant negative effects caused by the EHMT2 p.Ala1077Ser variant, our data and previously published data suggest that loss of EHMT2 function is probably more detrimental to cells than loss of EHMT1, explaining why individuals with alterations in *EHMT2* are very rare.

## INTRODUCTION

Dysregulation of the epigenetic machinery is associated with neurodevelopmental defects in humans. Kleefstra syndrome (KS) is a neurodevelopmental syndrome caused by heterozygous deletions at chromosome 9q34.3 that include the *EHMT1* gene (∼50%) or heterozygous intragenic *EHMT1* pathogenic variants (∼50%)^1,2^. In addition, loss-of-function mutations in the *KMT2C* (MLL3) gene cause a Kleefstra-like syndrome called Kleefstra syndrome-2 (KLEFS2)^3–5^. Moreover, although very rare, de novo mutations in epigenetic regulators genes *MBD5*, *MLL3*, *SMARCB1*, and *NR1I3* have been associated with intellectual disability disorders that fall into the Kleefstra syndrome’s phenotypic spectrum^3^.

EHMT1 (also known as GLP) and EHMT2 (also known as G9a) are highly homologous proteins that contain a SET domain that confers them the ability to mono- and dimethylate lysine 9 at histone H3 (H3K9)^6,7^. H3K9 methylation is finely regulated by several methyltransferases and demethylases and plays important roles in development, differentiation, and cancer^8^. EHMT1 and EHMT2 can form homo- or heterodimers through SET domain interactions, being heterodimers the most active configuration and the preferred form in cells^9^. Both Ehmt1-/- and Ehmt2-/- mice show severe developmental delays and early embryonic lethality that correlates with lower levels of H3k9me1 and me2^9,10^.

Despite the overlap in domains and functions of EHMT1 and EHMT2 and their relevance for development, patients with alterations in EHMT2 have never been reported. Here we report a Kleefstra-like syndrome case with a missense variant in the catalytic SET domain of EHMT2 that causes alanine 1077 change to serine reducing its affinity for the H3 tail and the activity of the enzyme by three- to five-fold. Results from our study allow defining this variant as pathogenic following the ACMG criteria^11^, since it appears the novo in a patient with disease and no family history (PS2), in vitro functional studies are supportive of a damaging effect (PS3), it is absent from controls in gnomAD (PM2) and the variant is located in a critical functional domain (PM1).

## MATERIALS AND METHODS

### Participants

Patients ND095 and ND120 were recruited by the undiagnosed rare diseases program SpainUDP^12^ at the Institute of Rare Diseases Research (IIER), Spanish National Institute of Health Carlos III (ISCIII). Peripheral blood samples and skin biopsies were collected from patients and their parents to perform trio-based whole-exome sequencing and establishment of fibroblast cultures respectively. Informed consents were signed by patient’s legal representatives. This research project was approved by the ISCIII Research Ethics Committee. Patient labels ND095 and ND120 correspond to an internal code of the SpainUDP program that is only available to the research team.

### Whole Exome sequencing

Whole exome sequencing and data analysis were performed in the probands and their unaffected parents. Genomic DNA was extracted from peripheral blood using the Qiagen QIAamp DNA kit. Whole Exome Sequencing (WES) libraries were prepared using the Nimblegen MedExome + ChrMit as enrichment kit and HS2000 v4, 2×100bp sequencing platform in ND095 family and Nextera Felx DNA Library Prep and Illumina NextSeq500 in ND120 family. Data analysis was performed using two different standardized protocols as previously described^12^. These included an in-house analysis and a parallel analysis using the Genomic Analysis module of the RD-Connect Genome-Phenome Analysis Platform (GPAP)^13^. All commonly identified rare variants were further analysed, checking all available scientific evidence through detailed searches in public databases (including Gene-Card, NCBI, UniProt, OMIM, Pubmed and ExAc). Sanger sequencing was performed to validate candidate variants.

### Fibroblast culture

Human dermal fibroblasts cultures were established from skin tissue samples. Briefly, fibroblasts were mechanically isolated by dissecting the dermal layer of the skin and the resulting fragments were incubated at 37°C in Dulbeccos’ modified Eagle’s medium (DMEM) containing 2% of fetal calf serum. Cells were expanded by incubation in 75 cm2 culture flasks at 37°C with 5% CO2 and 95% humidity in DMEM containing 10% fetal bovine serum.

### RNA-seq analysis

Total RNA was extracted from fibroblasts using the RNeasy mini kit from Qiagen. RNA-seq was performed at BGI Tech Solutions with two or three biological replicates per condition. Briefly, ribosomal RNAs were removed using a RNase H-based method and first-strand cDNA was generated using random hexamer-primed reverse transcription, followed by a second-strand cDNA synthesis with dUTP instead of dTTP, end repair, A addition and adaptor ligation. The U-labeled second-strand template was digested with Uracil-DNA-Glycosylase (UDG) and amplified by PCR. The resulting library was validated by quality control. The PCR products were then heat denatured and circularized by the splint oligo sequence to generate a single strand circle DNA followed by rolling circle replication to create DNA nanoballs (DNB) for sequencing on the MGI DNBSEQ platform. Raw sequencing data with adapter sequences or low-quality sequences was trimmed or filtered and examined by FastQC for basic quality controls. The sequencing analysis was carried out using Galaxy (https://usegalaxy.eu/). Paired reads were aligned to the human hg19 genome build using STAR^14^. Gene counts were calculated using HTseq-count^15^ and differential expression between patients and healthy fibroblast was interrogated using DEseq^16^. Differentially expressed genes were considered at an adjusted p-value <0.05.

For enrichment analysis in differentially expressed genes, we used GSEA preranked^17^ with genes ranked according to p-value corrected log2 of fold change. Interrogated gene sets were Gene Ontology Biological Process (GOBP) and transcription factor targets (TFT). In addition, we performed enrichment in GOBP terms in significantly upregulated or downregulated genes using Panther (https://pantherdb.org/)^18^. Scatterplots showing correlations were generated with ggplot2 in Galaxy. Pearson’s correlation and bubble plots were calculated using SRPlot^19^. Network representation of enriched GOBP terms was generated using Cytoscape Enrichment Map and AutoAnotate Apps^20^.

### Production of recombinant proteins

pET28a-LIC containing the catalytic domain of human EHMT2 (aa913-1193) was a gift from Cheryl Arrowsmith (Addgene plasmid # 25503; http://n2t.net/addgene:25503; RRID:Addgene_25503). Mutation p.Ala1077Ser was introduced using the QuikChange II XL Site-Directed Mutagenesis Kit from Agilent. Expression and purification of His-tagged EHMT2 catalytic domain WT and p.Ala1077Ser in bacteria has been previously described^21^. Recombinant H3 was expressed in *E.coli* and purified as previously described^22^

### Histone methyltransferase assays

Histone methyltransferase assays were performed using the fluorescence-based assay SAM Methyltransferase Assay SAMfluoro™ from G-Biosciences that detects the formation of resorufin over time as a result of the methyltransferase reaction. Assays were performed using 250, 500 or 1000 ng of His-tagged WT or p.Ala1077Ser EHMT2 catalytic domains and 2.5 µg of recombinant histone H3 following the manufacturer instructions. Cumulative reads were measured every minute for 45 minutes. Specific activity was calculated using a resorufufin standard.

### Histone peptide binding assays

10 µg of biotinylated histone 3 peptides aa 1 to 21 (Active Motif) unmodified, monomethylated at K9 or dimethylated at K9 were incubated overnight at 4°C with 5 ug of recombinant EHMT2 wild type or p.Ala1077Ser mutant catalytic domain in 300 µl of binding buffer 50 mM Tris pH 7.5, 150 mM NaCl, 0.05% NP-40 and 1 mM PMSF. After 1 hour incubation with Pierce streptavidin magnetic beads and extensive washing with binding buffer bound proteins were resolved by SDS-PAGE and stained with Coomassie blue. Gels were scanned using the Bio-Rad GelDoc Go Imaging System and bands quantified using ImageJ^23^.

### Protein modeling

PDB Protein Data Bank entry 5JJ0 (https://doi.org/10.2210/pdb5JJ0/pdb) corresponding to the catalytic domain of EHMT2 complexed with the histone peptide H3K9M and SAM was explored in 3D using PDB resources^24^.

### EpiSign assay

Methylation analysis was conducted using the clinically validated EpiSign assay, following previously established methods^25–28^. Methylated and unmethylated signal intensities generated from the EPIC array were imported into R 3.5.1 for normalization, background correction, and filtering. Beta values were then calculated as a measure of methylation level, ranging from 0 (no methylation) to 1 (complete methylation), and processed through the established support vector machine (SVM) classification algorithm for EpiSign disorders. The classifier utilized the EpiSign Knowledge Database, which consists of over 10,000 methylation profiles from reference disorder-specific and unaffected control cohorts, to generate disorder-specific methylation variant pathogenicity (MVP) scores. These MVP scores are a measure of prediction confidence for each disorder and range from 0 (discordant) to 1 (highly concordant). A positive classification typically generates MVP scores greater than 0.5. The final matched EpiSign result is generated using these scores, along with the assessment of hierarchical clustering and multidimensional scaling.

### Detection of histone modifications by mass spectrometry

Mass spectrometry was performed by Active Motif. Histones were acid extracted from a cell pellet containing 2.5×10^6^ cells, derivatized via propionylation, digested with trypsin, newly formed N-termini were propionylated as previously described^29^. Histones were extracted by incubating samples at room temperature for 1 hour in 0.2M sulfuric acid with intermittent vortexing. Histones were then precipitated by the addition of trichloroacetic acid (TCA) on ice, and recovered by centrifugation at 10,000 x g for 5 minutes at 4°C. The pellet was then washed once with 1mL cold acetone/0.1% HCl and twice with 100% acetone, and then air dried in a clean hood. The histones were propionylated by adding 1:3 v/v propionic anhydride/2-propanol and incrementally adding ammonium hydroxide to keep the pH around 8, and subsequently dried in a SpeedVac concentrator. The pellet was then resuspended in 100 mM ammonium bicarbonate and adjusted to pH 7-8 with ammonium hydroxide. The histones were then digested with trypsin, resuspended in 100mM ammonium bicarbonate overnight at 37°C, and dried in a SpeedVac concentrator. The pellet was resuspended in 100mM ammonium bicarbonate and propionylated a second time by adding 1:3 v/v propionic anhydride/2-propanol and incrementally adding ammonium hydroxide to keep the pH around 8, and subsequently dried in a SpeedVac concentrator. Histone peptides were resuspended in 50 µL of 0.1% TFA and 3 µl were injected with 3 technical replicates in a Thermo Scientific TSQ Quantum Ultra mass spectrometer coupled with an UltiMate 3000 Dionex nano-liquid chromatography system. The data was quantified using Skyline^30^, and represents the percent of each modification within the total pool of that amino acid residue.

## RESULTS

### Clinical characteristics of the proband overlap with Kleefstra syndrome

Proband ND095 was remitted to the undiagnosed rare diseases program SpainUDP^12^ after testing negative for candidate gene panels. The proband suffered from global developmental delay characterized by slow developing of gross motor milestones and expressive language delay, hypotonia without weakness, coarctation of the aorta, nephrocalcinosis and renal medullar cystic disease. At the craniofacial level, the proband had brachicephaly, plagiocephaly with very flat occiput, broad face and midface hypoplasia, synophrys, sparse medial eyebrows, epicanthus, lateral deviation of upslanted palpebral fissures, mildly everted lower eyelids, palpebral ptosis, anteverted nares, smooth philtrum and carp-like mouth. In addition, the proband had dental diastema, microdontia, persistent fetal fingertip pads and scoliosis (a complete clinical history can be provided upon request). From the clinical point of view, the patient’s phenotype overlaps significantly with the clinical characteristics of KS (Table 1).

**TABLE 1.** Anomalies found in patient ND095 compared to Kleefstra syndrome^1,2,44,46^.

| **Anomalies** | **ND095** | **Kleefstra** |
| --- | --- | --- |
| High birth weight | - | 8-50% |
| Microcephaly | - | 30-80% |
| Brachycephaly | + | 33% |
| Flat face | + | 16% |
| Hypertelorism | + | 55-70% |
| Arched eyebrows | + | 27% |
| Carp mouth | + | 77% |
| Brachydactyly | + | 16% |
| Synophrys | + | 55-80% |
| Epicanthal folds | + | 30-70% |
| Upslanting palpebral fissures | + | 30-70% |
| Midface hypoplasia | + | 55-100% |
| Anteverted nares | + | 40-80% |
| Protruding tongue | - | 40-60% |
| Ear anomalies | - | 45-80% |
| Dental anomalies | + | 10-15% |
| Hypotonia | + | 60-80% |
| Psychomotor delay/intellectual disability | + | 100% |
| Autism ASD | - | 30-75% |
| Short stature | - | 10-39% |
| Behavioral problems | stereotypy | 65-70% |
| Sleep disorder | - | 20-50% |
| Epilepsy | one episode | 20-50% |
| Brain imaging anomalies | partial empty sella turcica | 50-60% |
| Cardiovascular anomalies | + | 40-45% |
| Renal issues | + | 15-30% |
| Genital anomalies | + | 45-50% of males |
| Umbilical/inguinal hernia | + | 15-20% |
| Skeletal anomalies | + | 30-50% |
| Extremities | + | 70% |
| Hypoacusia | - | 20-30% |
| Ocular anomalies | - | 45% |
| Obesity | - | 30-40% |
| Respiratory complications | + | 5-14% |
| Gastro-esophageal reflux | - | 14-19% |
| Tracheomalacia | - | 5-11% |
| Recurrent infections | - | 26-64% |
| Speech anomalies | non-verbal | ~100% |

### Whole-exome sequencing reveals a VUS in the *EHMT2* gene

Whole-exome sequencing of patient ND095 and his parents carried out at SpainUDP revealed no detectable deleterious variants in the Kleefstra syndrome 1 (KS) causing gene *EHMT1*. In addition, no alterations were detected in *KMT2C,* which causes Kleefstra syndrome 2^5^. Similarly, no pathogenic variants were detected in chromatin-related genes *MBD5*, *MLL3*, *SMARCB1*, and *NR1I3* that have been previously associated with Kleefstra-like syndromes^3^. Surprisingly, the sequencing reveled a heterozygous de novo variant in the *EHMT2* gene (chr6:31848838C>A; NM_006709.5:c.3229G>T) that resulted in the amino acid change alanine 1077 to serine (p.Ala1077Ser). This variant was not present in gnomAD V4 genomes not exomes, neither reported in ClinVar. Several in silico predictors classified this variant as pathogenic (EIGEN, FATHMM, LRT, MutPred MVP, REVEL) while it was classified as variant of uncertain significance (VUS) by others (Mutation Taster, Mutation Assessor, SIFT, PROVEAN).

### The p.Ala1077Ser variant decreases the activity of EHMT2

Ala1077 is located in the catalytic SET domain of EHMT2, in a highly conserved region among human histone methyltransferases (Figure 1A). Ala1077 is, however, far from the AdoMet cofactor interacting amino acids (Figure 1A) and the presumed dimerization region (Figure 1B). The published EHMT2 catalytic domain structure in complex with the H3 tail^31^ shows that Ala1077 interacts with the H3 N-terminal tail residue threonine 6 suggesting that it might affect the catalytic activity of EHMT2 through changes in this interaction (Figure 1C). To test this possibility, we first evaluated the effects of the p.Ala1077Ser variant in the activity of the enzyme. We used an in vitro fluorescence-based assay that allows the detection of methyltransferase activity over time to test the activity of wild type and Ala1077Ser EHMT2 catalytic domains expressed and purified in *E.coli,* on recombinant histone H3. Figure 2A shows that the p.Ala1077Ser variant drastically reduces the methyltransferase activity of EHMT2 on histone H3 and the specific activity of the enzyme by five-fold (Figure 2B). Replication of the histone methyltransferase assay using different EHMT2 concentrations shows that the Ala1077Ser variant reduces the specific activity of the enzyme by three-to five-fold (Figure 2B, Supplementary Figure 1). Next, we evaluated if the p.Ala1077Ser variant affects the interaction of EHMT2 with histone H3 tail by testing the interaction of wild type EHMT2 and p.Ala1077Ser mutant with synthetic H3 peptides (aa 1-21) either unmethylated, mono or dimethylated at lysine 9, in an in vitro pull-down assay. Figure 2C and 2D show that the p.Ala1077Ser variant reduces de affinity of the catalytic domain for histone H3 tail. In addition, we detected a reduced affinity of the WT catalytic domain for the dimethylated peptide compared to unmethylated or monomethylated peptides. These results are in line with previous data that suggests loss of affinity of methyltransferases for its substrate once it is fully methylated^21,32^. This behavior was not observed in the mutant catalytic domain (Figure 2C and D).

**Figure 1.**
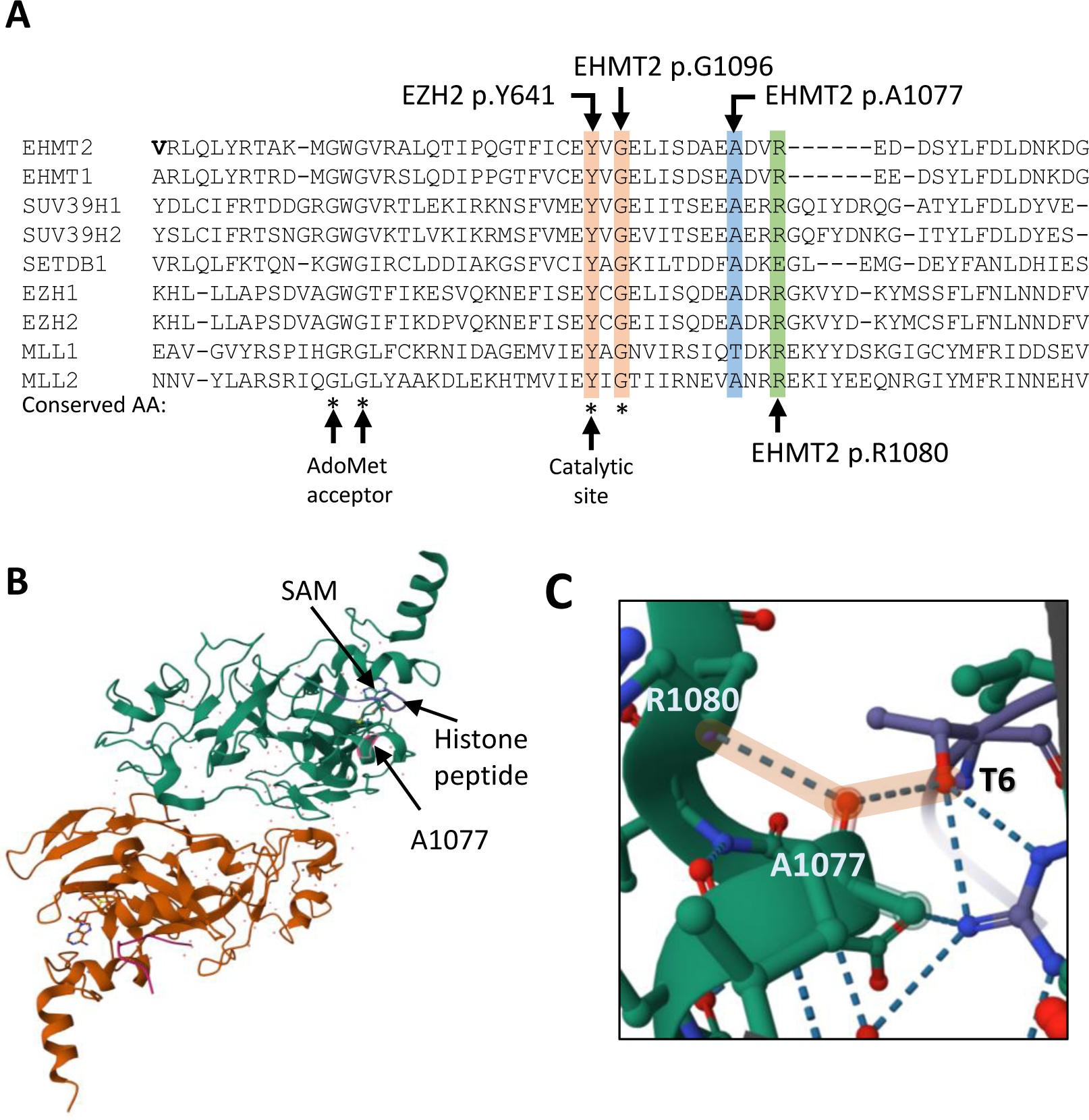
p.Ala1077 is located in the catalytic domain of EHMT2. A. Protein alignment of several human histone methyltranserases. p.Ala1077 and other amino acids described to confer gain of function are highlighted. Adapted from Kato S. et al^32^. B. Structure of EHMT2 SET-domain dimers with histone H3 (purple) and SAM (blue). C. Magnification shows that residue p.Ala1077 in EHMT2 stablishes hydrogen bonds (highlighted in orange) with histone tail residue T6 and EHMT2 residue p.Arg1088.

**Figure 2.**
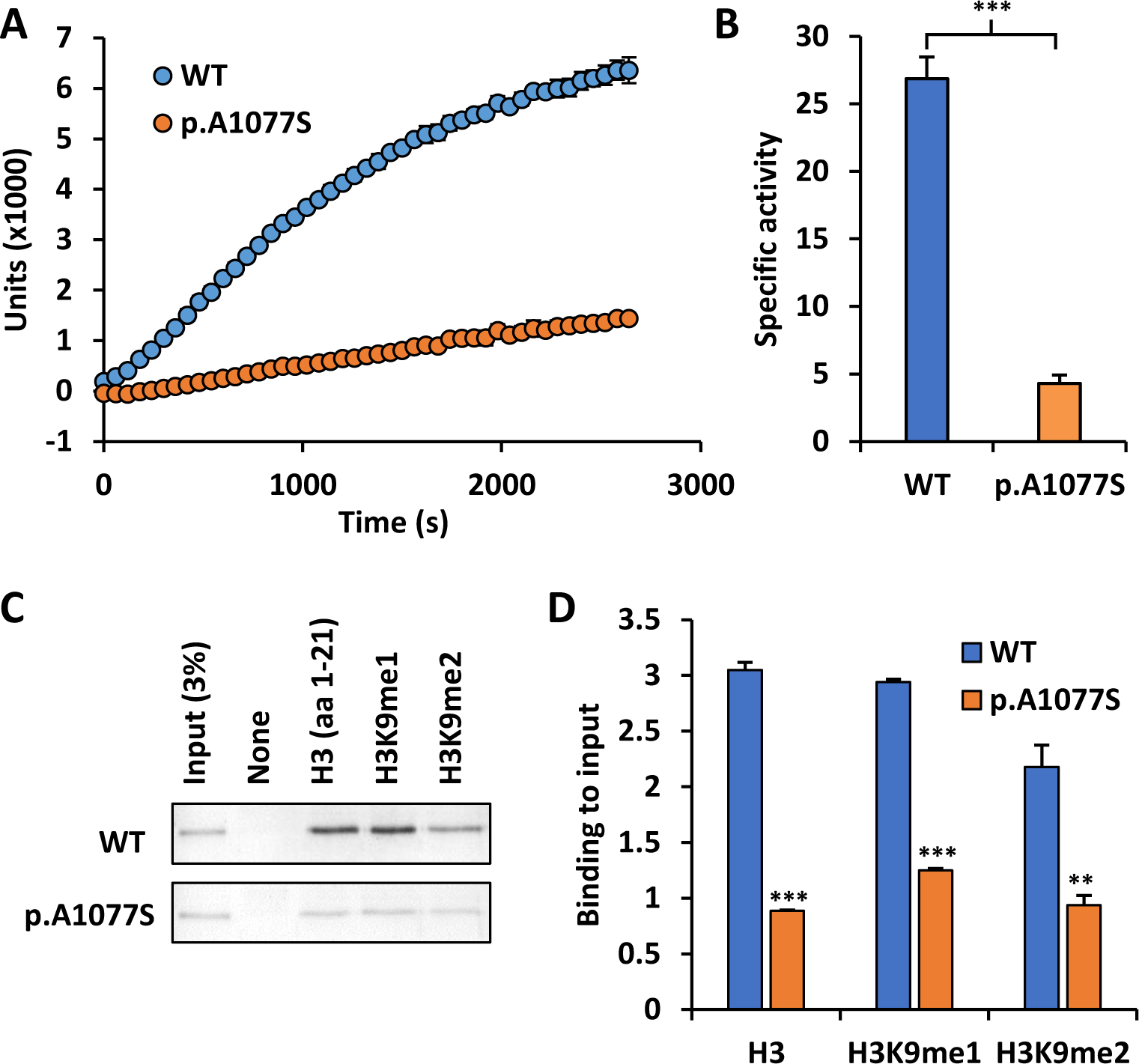
Molecular alterations in EHMT2 catalytic domain caused by the p.Ala1077Ser change. A. Measure of cumulative methyltransferase activity over time (seconds) using 250 ng of the catalytic domain of EHMT2 wild type (WT) or p.Ala1077Ser mutant and 2.5 µg of recombinant histone H3 as a substrate. Graph shows the mean and standard deviation of two replicates B. Specific activity in nmol/min/mg of WT and p.Ala1077Ser catalytic domains. Mean and standard deviation of two replicates is shown. C. Coomassie blue stained gel depicting the interaction of WT and Ala1077 catalytic domains with histone H3 peptides amino acids 1 to 21 unmodified (H3), monomethylated at lysine 9 (H3k9me1) and dimethylated at lysine 9 (H3k9me3) D. Quantification of WT and p.Ala1077Ser catalytic domains binding to peptides relative to input. Mean and standard deviation of three quantifications is shown. P-values for comparisons between WT and p.Ala1077Ser correspond to *<0.05, **<0.005 and ***<0.0005

### The Episign test classifies patient ND095 as Kleefstra syndrome

In order to investigate the overlap of patient ND095 with KS-specific DNA methylation signature, we compared the blood DNA methylation profile of patient ND095 with Kleefstra patients using version three of the test EpiSign^26^. This test, based in blood DNA methylation profiles, allows the diagnosis of over fifty different genetic diseases. Unsupervised hierarchical clustering of DNA methylation profiles shows that ND095 clusters with Kleefstra syndrome patients and clearly separates from healthy controls (Figure 3A). The computational model classified patient ND095 as KS with high confidence, generating a low score for other 56 syndromes (Figure 3C). However, the principal component analysis of DNA methylation showed that ND095 had a particular methylation profile that differs slightly from KS patients (Figure 3B). This data suggests the involvement of an *EHMT1*-related but different causal gene in the phenotype of patient ND095.

**Figure 3.**
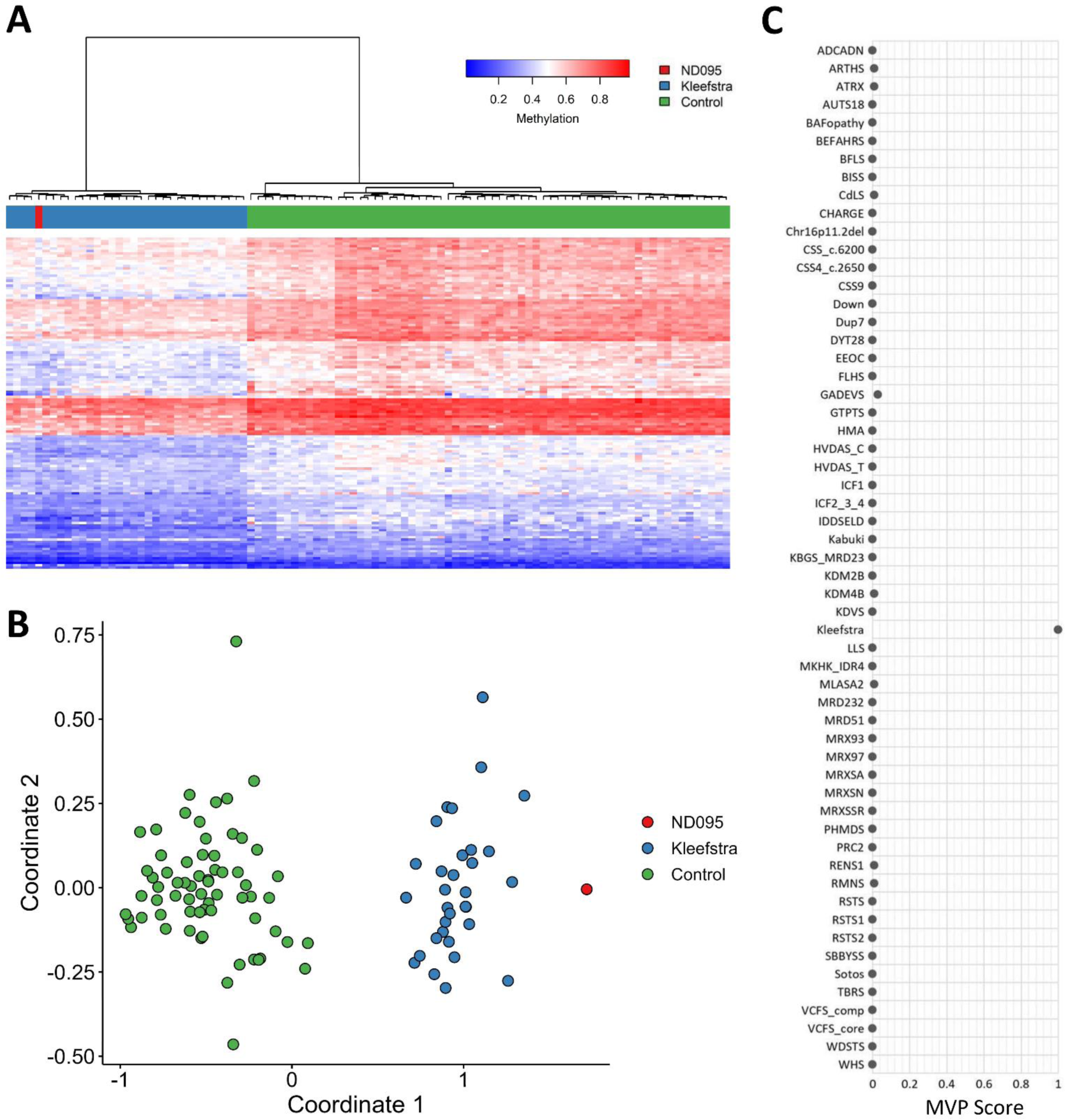
Results of the Episign test in patient ND095 A. Hierarchical clustering and B. multidimensional scaling plots indicate the case (red) has a DNA methylation profile similar to subjects with a confirmed Kleefstra episignature (blue) and distinct from controls (green). C. MVP score, a multi-class supervised classification system capable of discerning between multiple episignatures by generating a probability score for each episignature. The elevated score for Kleefstra syndrome indicates an episignature similar to the Kleefstra syndrome reference.

### Overlap of molecular signatures between patient ND095 and Kleefstra syndrome

Next, we established fibroblasts cultures from a healthy donor, the ND095 patient carrying the EHMT2 p.Ala1077Ser variant and a patient (ND120) recently diagnosed with KS through our program, who carries a frameshift variant in *EHMT1* (NM_024757.5:c.1881delT p.His620ThrfsTer12) and quantified histone modifications by mass spectrometry. Figure 4A shows that ND095 had decreased levels of mono-, di- and trimethylated H3K9 while levels of H3K27me3 remained unchanged. In correlation, we found an increase in unmodified and acetylated H3K9. Similar, but less dramatic effects, were found in KS patient ND120 (Figure 4A). These results are in agreement with a previous characterization of changes in histone modifications after knockdown for both EHMT2 and EHMT1^33^ and are compatible with the loss of EHMT2 activity in patient ND095.

**Figure 4.**
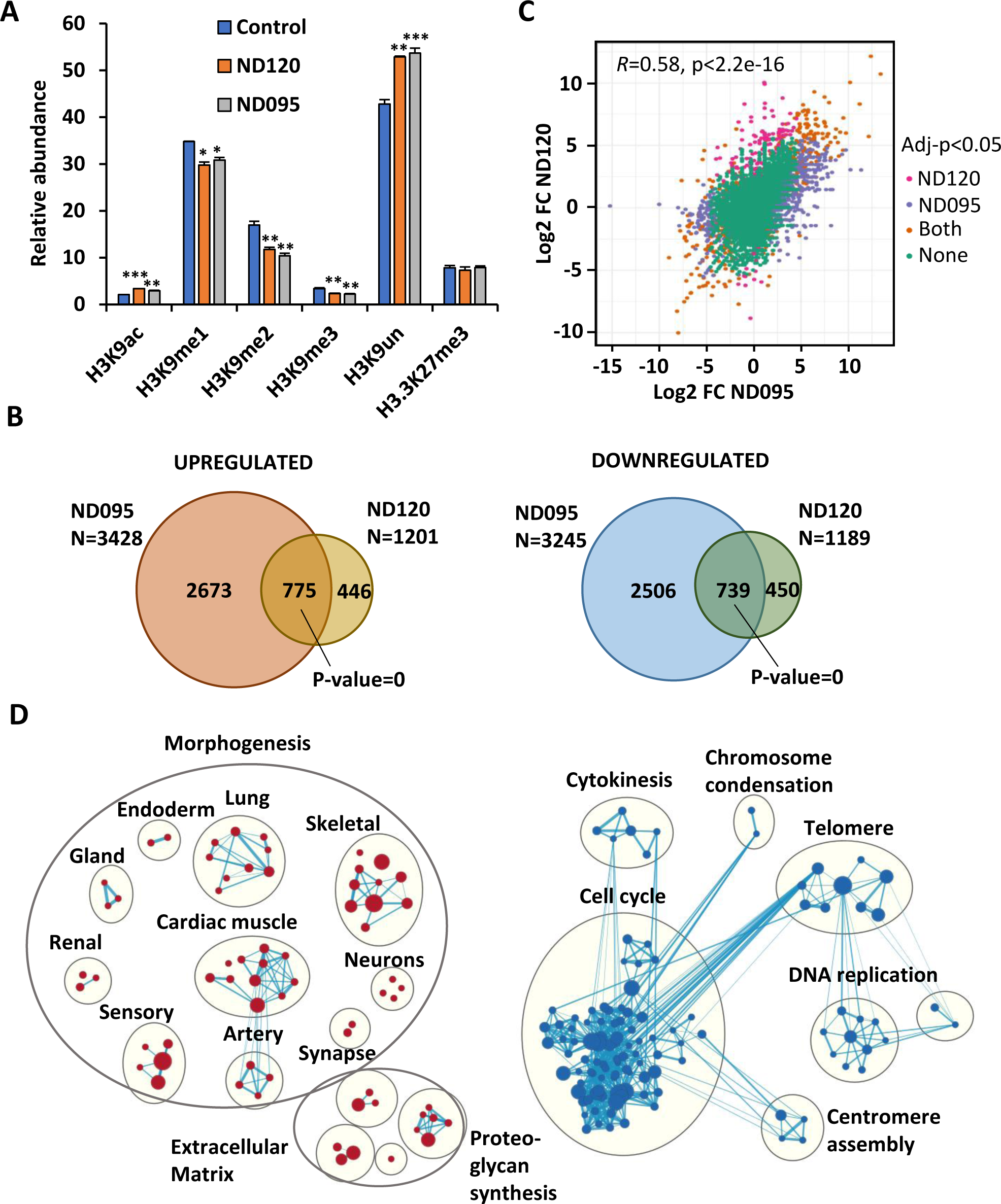
Transcriptomic and epigenetic analysis of fibroblast derived from a healthy control, a Kleefstra Syndrome patient (ND120) and the patient carrying the EHMT2 p.Ala1077Ser variant (ND095). A. Relative abundance of histone H3 modifications quantified by mass spectrometry in patient’s fibroblasts and a healthy donor. Bars show the mean and standard deviation of tripicates. Significant differences compared to control healthy fibroblast are indicated with asterisks according to P-values *<0.05, **<0.005 and ***<0.0005. B. Overlap of upregulated and downregulated genes found in ND095 or ND120 compared to the healthy control at adjusted p-value<0.05. C. Scatter plot showing correlation between expression fold change of all genes in ND120 or ND095 compared to control fibroblasts. Pearson’s correlation coefficient is indicated. Genes changing expression at an adjusted p-value<0.05 only in ND120 (pink), only in ND095 (purple), in both (orange) and not significantly changing expression (green) are shown. D. Network representation and clustering of GSEA results on GOBP terms at FDR<0.06. Red nodes correspond to terms enriched in genes upregulated in ND095 compared to control fibroblasts and blue nodes in downregulated genes.

Afterwards, we carried out RNA-seq in the fibroblasts cultures and identified differentially expressed genes (DEGs) in ND095 or ND120 fibroblasts compared to healthy fibroblasts. Among DEGs we found both upregulated and downregulated genes with a higher number of DEGs in ND095 compared to ND120 (Figure 4B). This correlates with the more severe effects in H3K9 methylation in ND095 found by mass spectrometry. Despite this difference there was a significant overlap in down and upregulated genes between ND095 and ND120 (Figure 4B) and a significant correlation comparing the fold change of expression of all genes (Figure 4C).

We next investigated the enrichment of GOBP terms in the DEGs using gene set enrichment analysis (GSEA). Results show a significant enrichment of genes involved in the morphogenesis of diverse tissues and organs derived from the three embryonic layers, and genes involved in the extracellular matrix in genes upregulated in ND095 compared to control fibroblasts (Figure 4D). Significant enrichment in morphogenesis genes in upregulated genes was also identified in patient ND120 (Supplementary Figure 2A) and in genes commonly upregulated in both patients (Supplementary Figure 2B). Genes downregulated in ND095 fibroblasts compared to healthy fibroblasts were mainly involved in cell cycle (Figure 4D). Additionally, a significant enrichment of E2F transcription factors binding sites was found around the transcriptional start site of genes downregulated in ND095 fibroblasts (Supplementary Figure 3A). Moreover, the expression of several E2F factors was significantly downregulated in ND095 fibroblasts (Supplementary Figure 3B). However, in ND120 we could not find enrichment of cell cycle related genes, nor genes bound by E2F transcription factors, in downregulated genes or downregulated E2F factors.

### Changes in *EHTM1* and *EHMT2* expression in patients’ fibroblasts

Interestingly, the expression of *EHMT2* was significantly downregulated in ND095 compared to healthy control fibroblasts suggesting that p.Ala1077Ser has consequences for *EHMT2* mRNA expression (Supplementary Figure 4A). Despite this, the reads covering the NM_006709.5:c.3229G>T region showed a balance of 50% WT and 50% mutant transcripts (Supplementary Figure 4B). In patient ND120, we detected lower mRNA expression of both *EHMT1* and *EHMT2* (Supplementary Figure 4A). Reads in the EHMT1 NM_024757.5:c.1881delT region were predominantly WT suggesting that the T deletion affects the levels of RNA likely by nonsense-mediated decay (Supplementary Figure 4B).

## DISCUSSION

We describe here a patient with a de novo pathogenic missense variant in the *EHMT2* gene that causes a Kleefstra-like syndrome phenotype. This missense variant causes the change of alanine 1077 to serine in the catalytic SET domain of EHMT2 reducing its affinity for histone H3 tail and catalytic activity by three-to five-fold. Interestingly, the presence of p.Ala1077Ser change not only reduces the affinity of the catalytic domain for histone H3 but also abrogates the specificity of H3 recognition dependent on its modification. Loss of affinity of EHMT2 catalytic domain for the H3 tail once is dimethylated might favor the recruitment of other methyltransferases able to trimethylate histone H3. This recruitment might be blocked by the low but persistent binding of the p.Ala1077Ser mutant domain to H3K9me2.

Although the changes in gene expression and H3K9 methylation detected in patients’ fibroblasts are more dramatic in ND095 than ND120 there is a significant correlation in changes of gene expression between both patients. In both cases, there is an upregulation of genes involved in the morphogenesis of diverse organs and tissues, suggesting that EHMT1/2 play a role in preventing the expression of genes belonging to alternative lineages. In accordance, EHMT1/2 have been previously involved in the silencing of alternative lineage genes during hematopoietic differentiation^34,35^.

Regarding downregulated genes, a significant enrichment in cell cycle genes and E2F targets was only observed in patient ND095. Similarly, the depletion of EHTM2 in myoblasts has been previously shown to affect cell cycle through downregulation of E2F target genes^36^. This function has been suggested to be independent of EHMT2 methyltransferase activity and through its association with the E2F1/PCAF complex. Also, a downregulation of cell cycle genes and E2F-regulated factors has been described in KS induced pluripotent stem cells-derived neurons when compared to their healthy counterparts^37^.

It is surprising that while patients with alterations in *EHMT1* are known since 2004, reported as a syndrome caused by subtelomeric deletions of chromosome 9q^38^, individuals with genetic alterations in *EHMT2* have not yet been described. In contrast, EHMT2 was reported to be a histone methyltransferase before EHMT1^6^, and has been more extensively studied. Both proteins are highly similar, widely expressed in human tissues and together with the H3K9 methylation mark have been described to play critical roles in differentiation and development. Despite this, several studies suggest that EHMT2 and EHMT1 might have different functions being EHMT2 more relevant for maintaining the cellular levels of H3K9 methylation^9,39,40^. Although both EHMT1 and EHMT2 can methylate histones in vitro on their own, heterodimers between both proteins are needed to display maximum methylation activity in vivo^9^. Dimer formation-competent but enzymatically inactive mutant Ehmt1 but not Ehmt2 can rescue specific knock out (KO) mouse embryonic stem cells (mESCs) from defective H3K9 methylation, suggesting that the catalytic activity of EHMT1 but not EHMT2 is dispensable for the complex H3K9 methyltransferase activity in vivo^39^. In addition, it has been suggested that some of the defects observed after EHMT1 depletion could be due to altered EHMT2 activity. The Ehmt1 KO in mESCs results in reduced Ehmt2 expression likely due to the impossibility to form heterodimers, while the Ehmt2 KO had no effects in Ehmt1 expression^9,40^. In addition, KS causing EHMT1 variants p.Cys1073Tyr and p.Arg1197Trp, which have been reported to have both defective in vitro activity and interaction with EHMT2, did not rescue the levels of H3K9 methylation nor restored EHMT2 levels when overexpressed in Ehmt1 KO mESCs^40^. Therefore, it is likely that the EHMT1 truncations observed in Kleefstra syndrome patients affect the catalytic capacity of EHMT2 through alterations in heterodimer formation. These observations agree with the reduced levels of EHMT2 expression found in the KS patient ND120.

Our H3K9 methylation and gene expression data suggests that EHMT2 loss-of-function has more dramatic consequences regarding loss of H3K9 methylation and altered gene expression than loss of EHMT1 function, despite the fact that the p.Ala1077Ser change still retains some catalytic activity. This data agrees with the previously discussed more relevant role for EHMT2 in controlling the levels of H3K9 methylation than EHMT1. In this context, it is possible that loss of EHMT2 function might be more detrimental to cells than loss of EHMT1 function explaining why inactivating mutations in EHMT2 compatible with life are very rare. In agreement, mouse models have shown that the phenotypes of Ehmt1−/−embryos were mostly identical to those of Ehmt2−/−embryos, both leading to embryonic lethality^9,10^. Therefore, it is expected that complete loss of any of these proteins in humans results in severe developmental defects and are not compatible with life. In accordance with the human phenotype, both Ehmt1+/- and Ehmt2+/- mice are viable and recapitulate certain neurological traits observed in KS patients^41^.

However, the interpretation of the effects of missense variants versus truncating variants is not straightforward. Indeed, according to GnomAD^42^ V4 *EHMT1* is much more intolerant to loss-of-function variants than *EHMT2* (pLI=1 and LEUOF = 0.1 90%(0.07 - 0.16) vs. pLI = 0.86 and LEUOF = 0.39 90%(0.31 - 0.49), respectively), while EHMT2 is more intolerant to missense variants (Z = 1.98 and o/e = 0.87 (0.84 – 0.91) for *EHMT1* vs. Z = 4.63 and o/e = 0.68 90% (0.64 - 0.71) for *EHMT2*). While tools like DOMINO^43^, which predicts pathogenicity of genes based on their properties rather than variants suggests a very likely dominant inheritance for *EHMT2* in Mendelian disorders (probability: 0.8349), GnomAD probabilities suggest that monoallelic inactivating mutations could be tolerated. If this were the case, the pathogenicity of the monoallelic alteration in patient ND095 could be explained by potential dominant negative effects caused by the EHMT2 p.Ala1077Ser variant, such as sequestering EHMT1 into less active heterodimeric forms, that could lead to more severe effects than just the loss of one allele. Consequently, only a limited number of *EHMT2* non-truncating variants could act via this mechanism which could also explain the rarity of cases. Importantly, a clear correlation between *EHMT1* pathogenic variants and phenotype severity has not been yet established for KS^44^, reflecting the complexity of variant interpretation. In addition to deletions in chromosome 9q34.3 that eliminate one *EHMT1* copy, most common forms of *EHMT1* loss-of-function reported in KS patients is single nucleotide changes that cause frameshifts. Missense mutations are rarer and only a couple have been properly validated^40^. Larger efforts would be needed to evaluate the effects of *EHMT1* and *EHMT2* missense variants and their consequences for complex function and disease severity.

Regarding diagnostic perspectives for patients with *EHMT2* alterations, we found a DNA methylation signature for patient ND095 that resembles KS patients, likely due to the formation of one complex between EHMT1 and EHMT2. This shared, but slightly different, DNA methylation profile has been observed also for other disorders of the same protein complex such as BAFopathies^45^. However, the increased number of patients with rare diseases tested for DNA methylation profiles has improved the resolution and specificity of the episignatures ranging from altered protein complexes to genes, protein domains, and even single nucleotides^28^. Therefore, it is possible that a specific DNA methylation episignature for EHMT2 loss-of-function different from patients with KS can be established in the future after the identification and profiling of more patients with pathogenic *EHMT2* alterations.

## Data Availability

All data produced in the present study are available upon reasonable request to the authors

## ACKNOWLEDGEMENTS

This work was funded with project PID2021-128087OB-I00 by MCIN /AEI /10.13039/501100011033 / FEDER, UE to M.J.B, and AESI PT20CIII/00009 (ISCIII Platform of Biobanks and Biomodels PT-20). Funding was also partially provided by the Genome Canada and the Ontario Genomics Institute Genomics Applications Partnership Program Grant (OGI-188). This study used tools provided through the RD-Connect GPAP, which received funding originally from the European Union Seventh Framework Programme (FP7/2007-2013) under grant agreement No. 305444. We also thank the Bioinformatics Unit at ISCIII (I. Cuesta and S. Monzon) for their help in exome analysis, and A. Krepischi and L. Machado for discussions and S. Hortelano for advice on graphs. We thank the patients’ association Kleefstra España for their support, the National Biobank of Rare Diseases (BioNER) for helping with patient samples and the entire SpainUDP consortium for their collaboration in this program. The authors also would like to express their gratitude to the patients, their families and their physicians for their active participation in SpainUDP. The authors appreciate the support of the UDNI for international data sharing.

