## Supplementary Figures for "EHMT2 LOSS-OF-FUNCTION ALTERATIONS CAUSE A KLEEFSTRA-LIKE SYNDROME"

**Supplementary Figure 1**. Measure of cumulative methyltransferase activity along time (seconds) using 500 ng (A) or 1000 ng (C) of the catalytic domain of EHMT2 wild type (WT) or p.Ala1077Ser mutant and 2.5 µg of recombinant histone H3 as a substrate. B Specific activity in nmol/min/mg of WT and p.Ala1077Ser catalytic domains calculated using the assay conditions described in A. D. Specific activity in nmol/min/mg of WT and p.Ala1077Ser catalytic domains calculated using the assay conditions described in C. Mean and standard deviation of three replicates is shown in all graphs. P-values correspond to **<0.005 and ***<0.0005

**Supplementary Figure 2.** A. Network representation and clustering of GSEA results on GOBP terms enriched at p-value<0.05 in genes upregulated in ND120 compared to control fibroblasts. B. Bubble plot of GOBP enrichments found in genes commonly upregulated in ND120 and ND095 compared to control healthy fibroblasts.

**Supplementary Figure 3.** A. Normalized enrichment scores (NES) on transcription factor binding sites obtained by GSEA in DEGs between ND095 and healthy fibroblasts at a FWER p-value<0.05. NES for gene sets consisting of genes having at least one occurrence of the Y axis indicated TRANSFAC v7.4 motif in regions -/+ 2Kb around the TSS are shown. Negative enrichments denote enrichments in genes downregulated in ND095. P-value corresponds to FWER p-value and counts refer to the number of genes in the leading edge of each gene set. B. Heatmap of the mRNA expression of E2F transcription factors in the indicated sample. Asterisk denotes genes that were significantly downregulated (FDR<0.05) in ND095. No significant change in gene expression was detected in ND120.

**Supplementary Figure 4.** A. EHMT1 and EHMT2 mRNA levels in control healthy fibroblasts, fibroblast derived from a Kleefstra syndrome patient (ND120) and the patient carrying the EHMT2 p.Ala1077Ser variant (ND095). B. Percentage of wild type (WT) and mutant (MUT) reads detected in the RNA-seq from fibroblasts in EHMT2 in patient ND095 (total reads=98) and EHMT1 in patient ND120 (total reads=65). P-values *<0.05, **<0.005 and ***<0.0005

### Supplementary Figure 1

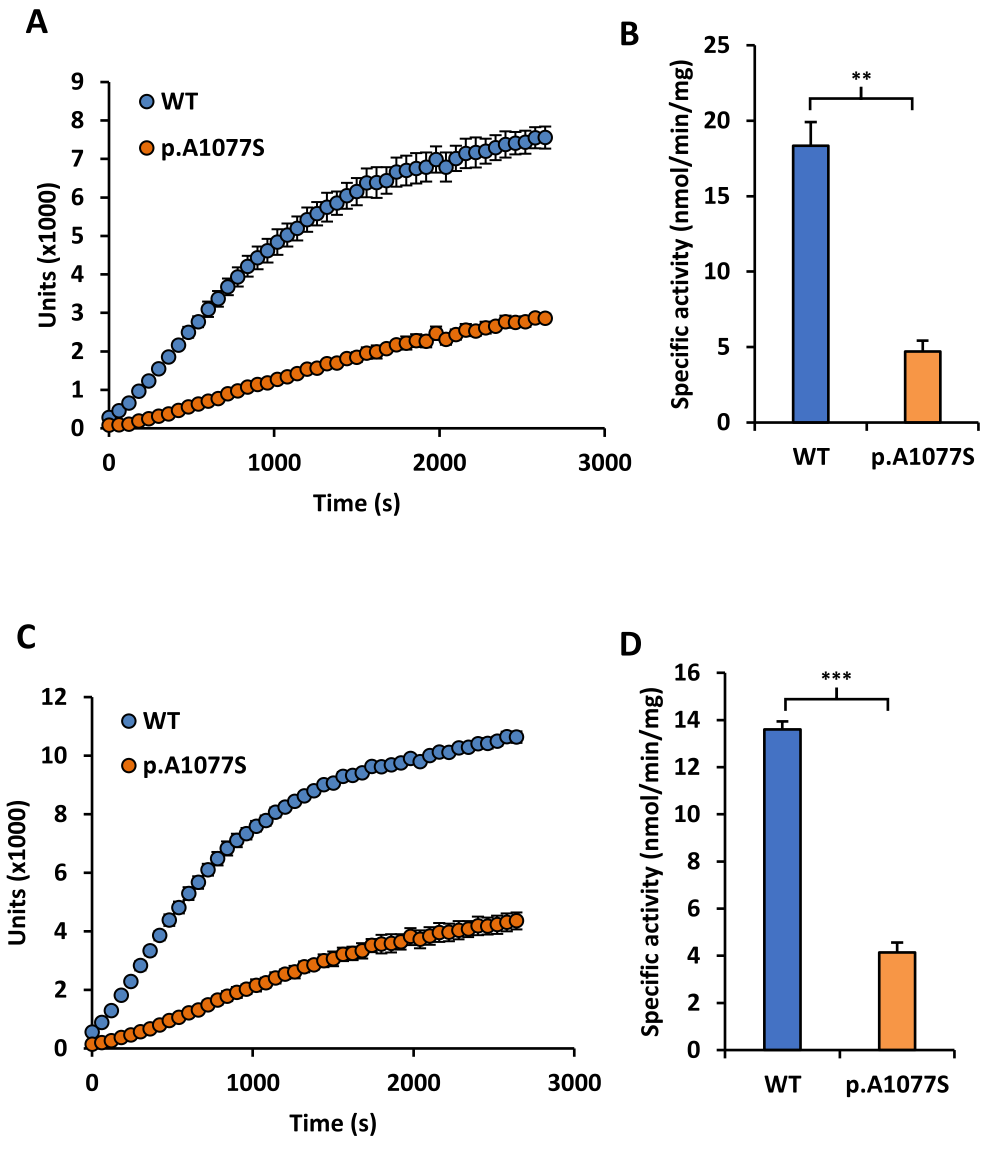

### Supplementary Figure 2

A

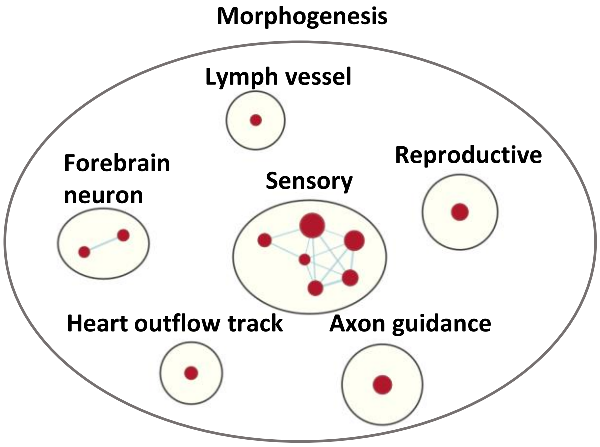


B

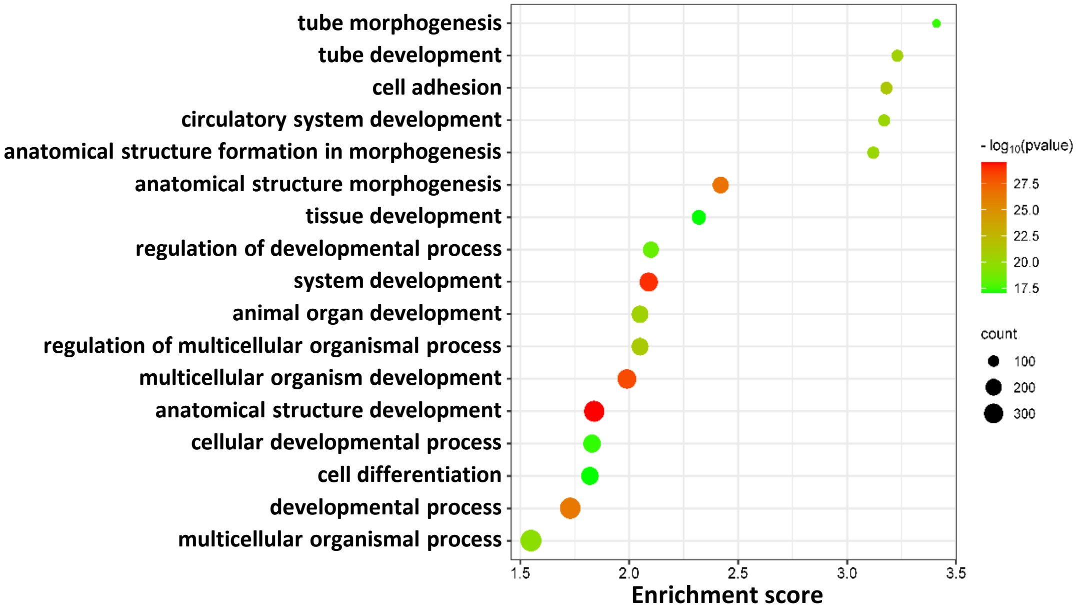

#### Supplementary Figure 3

A

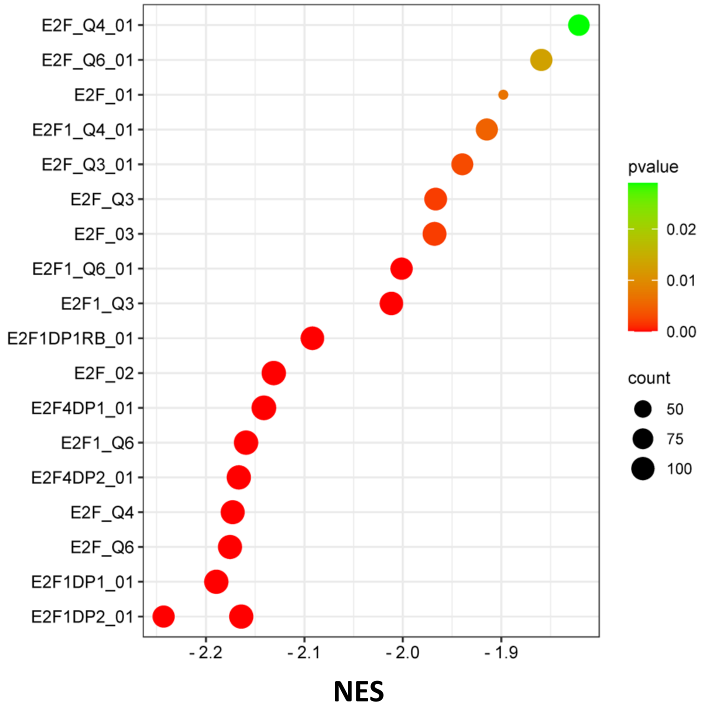


B

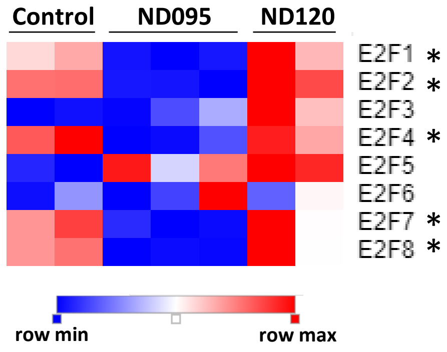

### Supplementary Figure 4

A

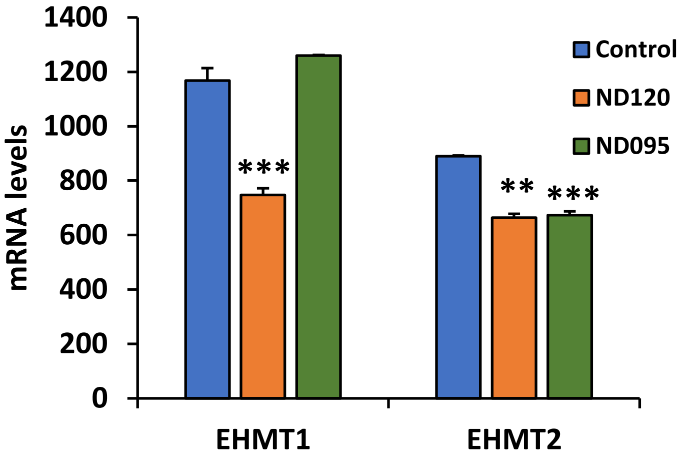


B

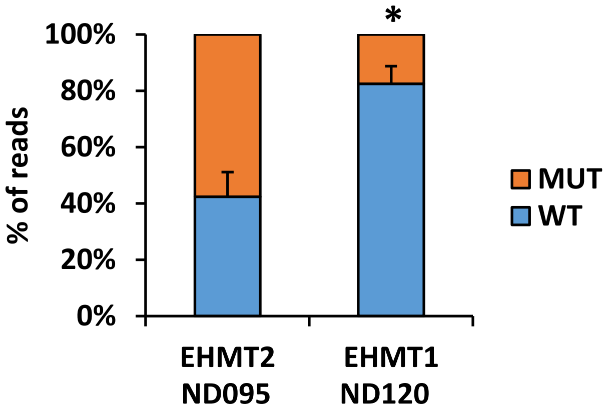
